# Clinical efficacy observation and proteomics of moxibustion in the treatment of menopausal obesity

**DOI:** 10.64898/2026.02.02.26345420

**Authors:** Benlu Yu, Zhongyu Zhou, Yunbao Zhu

**Affiliations:** Department of Acupuncture, Hubei Provincial Hospital of Traditional Chinese Medicine, No. 4 Garden Hill, Liangdao Street, Wuchang District, Wuhan 430061, Hubei, China; The First Clinical College of Hubei University of Traditional Chinese Medicine, No. 01 Tanhualin, Wuchang District, Wuhan 430060, Hubei, China

**Keywords:** Menopausal obesity, Moxibustion, Proteomics, HNRNPC

## Abstract

**Background:** Menopausal obesity is a type of obesity in women during menopause where the decline of ovarian function and the decrease of estrogen levels lead to an imbalance between energy intake and consumption in the body, resulting in fat accumulation and weight gain. Moxibustion, as a green therapy of non-interventional external treatment that prevents and treats diseases through thermal stimulation of relevant acupoints, has been widely used in clinical practice because of its simplicity, convenience, effectiveness, low price and high compliance.

**Purpose:** To clarify the pathogenesis of menopausal obesity and the biological mechanism of moxibustion treatment for menopausal obesity.

**Methods:** We selected 9 plasma samples from menopausal obese patients before and after moxibustion treatment, as well as 9 plasma samples from the healthy control group. After sample mixing and replication, DIA quantitative proteomics analysis was used to screen out differentially expressed proteins, and bioinformatics analysis was conducted.

**Results:** The plasma proteomic analysis revealed a significant increase in the protein expression levels of APOC2 and PZP in menopausal obesity patients. These differential proteins primarily participate in biological regulation, cell metabolism, and reproductive development processes. Their biological processes and molecular functions are mainly associated with enzyme inhibitor activity, calcium-dependent protein binding, lipid localization, and plasma lipoprotein particle assembly. The pathogenesis of menopause obesity is linked to the accumulation of visceral fat resulting from changes in sex hormone levels and reduced energy consumption following the decline of female ovarian function. Following moxibustion treatment, there was a notable down-regulation in the plasma levels of sialoglycoprotein receptor 2 (ASGR2), membranin A1 (ANXA1), and human heterogeneous nuclear ribonucleoprotein C (HNRNPC) among menopausal obesity patients. Their biological processes and molecular functions were primarily concentrated on intracellular hagy, nucleic acid binding, tissue regeneration, and neutrophil clearance.

**Conclusion:** The mechanism underlying moxibustion’s effectiveness in treating menopausal obesity may involve down-regulating HNRNPC expression, activating the PI3K/Akt/mTOR autophagy signaling pathway, regulating hormone levels to delay ovarian aging thereby improving lipid metabolism.

## Introduction

Menopause is a phase when the levels of FSH rise^[1]^. During this life stage, women are prone to a fast rise in visceral fat due to neuroendocrine dysfunction, increased glucocorticoid release, reduced metabolic rate, and other factors^[2–3]^. According to the Global Burden of Disease Obesity Collaborators, 603.7 million persons worldwide are obese^[4]^, with the prevalence of overweight or obesity ranging from 39% to 49%^[5]^. In addition to aggravate the risk of breast cancer, endometrial cancer, colon cancer, type 2 diabetes, and cardiovascular disease^[6]^, menopausal obesity also causes mental health conditions like depression, unjustified stigma, and schizophrenia and moves up health care expenses in nations all over the world^[7–9]^. A meta-analysis found that obesity was related with a 25% to 30% relative increase in breast cancer-specific and overall mortality in menopausal women^[10]^.

The primary interventions for obesity currently consist of lifestyle modification, pharmacological treatment, bariatric surgery, and traditional Chinese medicine therapy^[11]^. A simple diet and exercise regimen is sometimes challenging to maintain and prone to relapse^[12]^. Western pharmacological agents for weight reduction require explicit indications^[13]^, and bariatric surgery for obesity has an elevated risk of cardiovascular disease and death, in addition to being expensive^[14]^. Therefore, the pursuit of safe and effective medicines for the prevention and management of obesity during menopause has generated heightened concern.

Moxibustion has a high value of health economics, not only can save the patient’s time, energy and economic costs of the clinic, but also through the regulation of adipokines, hormone levels and other disease-causing factors to achieve the clinical efficacy of the first level of prevention, reducing the obesity and menopause to the individual and the community brought about by the double burden of the medical resources to reduce the consumption. A number of studies have shown that moxibustion can regulate hormone levels through the neuroendocrine-immune system, alleviate negative emotions such as anxiety and depression, delay ovarian aging, and correct glucose and lipid metabolism disorders, thus achieving the dual purpose of alleviating menopausal symptoms and reducing obesity^[15–18]^.

Proteomics is the analysis of the whole array of proteins expressed within a cell, tissue, or organism, characterized by large scale and high throughput. Through proteomics technology, proteins in biological samples and their corresponding modification sites may be identified and quantified extensively, enabling the extraction of useful insights and principles from extensive data analysis, so establishing a basis for the investigation of biological functioning systems. Based on previous clinical studies, DIA quantitative proteomics technology was used to detect plasma proteins of pre-menopausal and post-menopausal obese patients treated by moxibustion, and differentiated proteins were identified to ascertain the attributes of differentially expressed proteins and the activation of associated cellular processes, providing a theoretical basis for in-depth research on the pathogenesis of menopausal obesity and the biological mechanism of moxibustion for the treatment of menopausal obesity.

## Materials and methods

### Patients and samples

This study was conducted on voluntary subjects from the menopausal obese population of Hubei Provincial Hospital of Traditional Chinese Medicine. And the protocol of this study was registered in the International Clinical Trials Registry (Clinical Trials.gov) on 27 June 2021 with the protocol registration number NCT04943705^[19]^. The study design adhered to the Declaration of Helsinki criteria, complied with medical ethics regulations, and received approval from the Ethics Committee of Hubei Hospital of Traditional Chinese Medicine(HBZY2021-C17-01). Comprehensive information on the experiment will be conveyed to participants, and signed informed consent will be acquired from each subject following the initial screening. Patients (n = 9) who fulfil both menopause and obesity diagnostic criteria and healthy controls (n = 9)^[20–21]^ are recruited. Other inclusion criteria included those who had not used any hormonal and medicines associated with weight loss in the past three months and informed consent will be obtained from the study participants prior to study commencement. Exclusion criteria were unexplained vaginal hemorrhage or atypical menopause. Patients who have utilized hormone replacement therapy or other medications pertinent to this condition within the preceding three months, complicated by severe organic diseases, endocrine disorders, cardiovascular, hepatic, renal, and respiratory diseases, secondary to hypothalamic lesions, hypothyroidism, glucocorticoid therapy, and other factors leading to obesity. Concomitant with organic uterine abnormalities, including polycystic ovary syndrome, malignant tumors, severe cervical erosion, and complete hysterectomy, among others. Patients with contraindications to moxibustion, including moxa sensitivity, dermatological allergies, cicatricial constitution, and other severe skin conditions, will also be excluded. Subsequently, venous blood was collected from menopausal obesity before and after “moxibustion for harmonization of Yin and Yang” treatment and from healthy controls in a fasting state, and all subjects remained fasted for 12 hours before blood collection. The collected blood samples were firstly collected into the collection vessel with EDTA anticoagulant added, and gently reversed to make the anticoagulant and blood fully mixed. Then, centrifuge (Eppendorf-MiniSpin) was centrifuged at 3000r/min for 15min, and supernant was taken into the new screw-cap centrifuge tube, and plasma samples were divided into 200uL/ tube. Finally, it is rapidly frozen using liquid nitrogen, maintained in a refrigerator at -80 °C, and delivered on dry ice.

### Study interventions

The treatment was conducted by licensed Chinese medicine doctors with over 5 years of experience in clinical practice. In“Moxibustion for harmonization of Yin and Yang” group (Group A), moxibustion was applied within a circular area with a radius of 8-10 cm centered around the Mingmen point (DU04). In the second week, the treatment was administered similarly around the Shenque point (CV8), with treatments alternating weekly. Each session lasted 60 minutes, with one session per week, culminating in a total of 12 sessions over a 12-week period. Gentle moxibustion group (Group B) prescriptions for postmenopausal diseases and obesity, as well as clinical expert argumentation, were utilized to select the moxibustion points Zhongwan(RN12), Guanyuan(RN4) and Sanyinjiao(SP6) based on the textbook “Acupuncture and Moxibustion Therapy” published by China Traditional Chinese Medicine Publishing House in the 14th Five-Year Plan (Figure 1).

**Figure 1.**
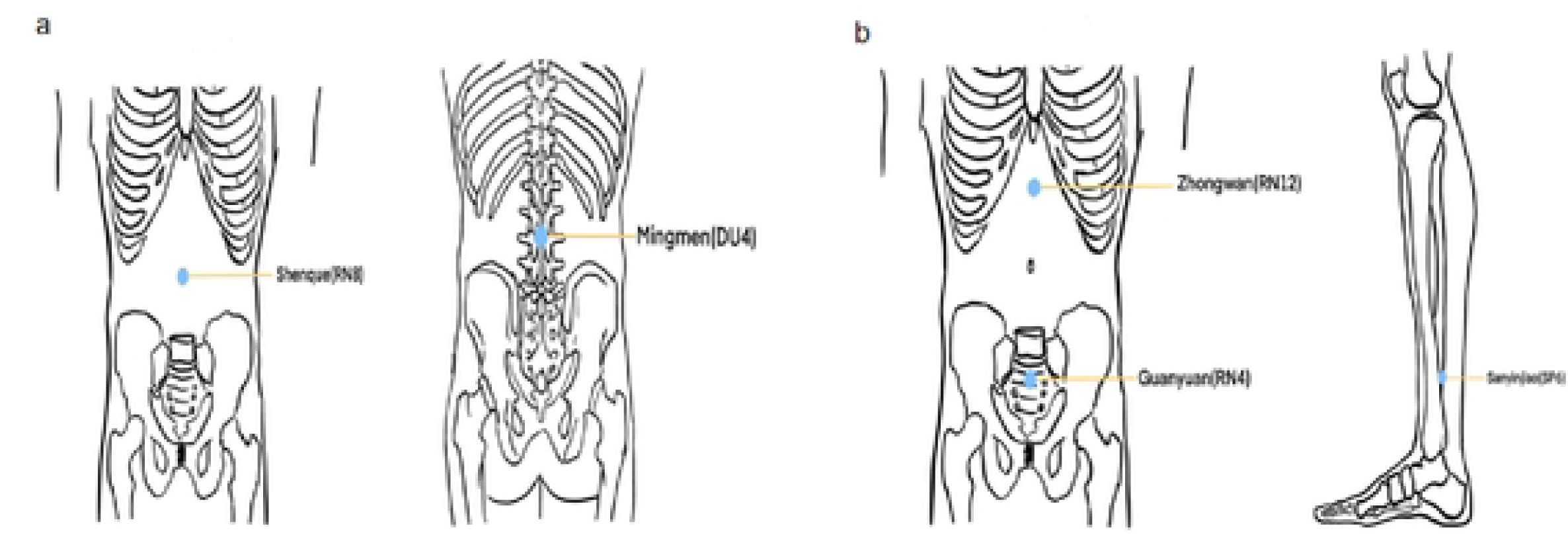
Acupoints selection of moxibustion. a: Locations of acupoints in the “Moxibustion for harmonization of Yin and Yang” group. b:Locations of acupoints in the “Moxibustion for harmonization of Yin and Yang” group

The treatment equipment included a single-hole moxibustion box (specifications: top 85mm*71mm, bottom 97mm*86mm, height 88mm, Li Shizhen Moxibustion Group Qichun Industry Qichun Co., LTD., product standard: Q/GJQA 010) and moxa sticks (specifications: 25g/piece, Hubei Li Shizhen Chinese Medicine Slices Co., LTD., Sinomedical license: Z20163081). One end of the moxa stick was ignited and placed in the moxibustion box so that it hung approximately 2-3cm above the skin. The local skin redness and perceived warmth of the patient were used as indicators of treatment intensity. Each point was subjected to treatment for 20 minutes every alternate day, three times weekly, throughout a duration of twelve weeks.

#### Assessment of menopausal obesity

All participants will fill out a questionnaire during the screening phase about demographic information, including gender, age, nationality, educational attainment, employment, marital status, height, weight, and relevant medical conditions, such as onset and duration. Each participant will have a clinical assessment of outcome measurements at two intervals: prior to treatment (screening period, initial visit) and following treatment (12th week).Waist circumference is the most straightforward and practical metric for assessing obesity. The pace of waistline decrease will be compared to the baseline, with assessments completed at weeks 0 and 12. Furthermore, the modified Kupperman score assesses the clinical manifestations of menopause before to and during therapy. The total score runs from 0 (minimum) to 63 (highest), with lower values signifying milder clinical symptoms in the patient.

#### Protein extraction and enzymolysis

A reaction solution (1% SDC/100 mM Tris-HCl pH 8.5/10 mM TCEP/40 mM CAA) was introduced to the samples and incubated at 60 °C for 1 hour to achieve simultaneous protein denaturation, reduction, and alkylation. Following dilution with an equivalent amount of ultrapure water, trypsin was introduced at a mass ratio of 1:50 enzyme to protein and incubated at 37 ℃ overnight with agitation for digestion. On the subsequent day, TFA was introduced to halt the digestion, and the supernatant was centrifuged at 16,000 g and desalted on a homemade SDB desalting column. The supernatant was desalted by centrifugation at 16000 g and frozen at -20 ℃ for use.

#### Mass Spectrometry

Mass spectrometry studies were conducted with a Bruker ion mobility-quadrupole-time-of-flight mass spectrometer, the timsTOF Pro. Sample injection and separation were conducted using an UltiMate 3000 RSLCnano liquid chromatograph, which was directly linked to the mass spectrometer. Peptide samples were extracted by an autosampler and affixed to a Trap column (75 μm × 20 mm, 2 μm particle size, 100 Å pore size, Thermo), then eluted to an analytical column (75 μ m × 250 mm, 1.6 μ m particle size, 100 Å pore size, ionopticks) for separation. An analytical gradient was created using two mobile phases: mobile phase A consisting of 0.1% formic acid in H_2_O and mobile phase B including 0.1% formic acid in ACN, for a duration of 60 minutes. The liquid phase flow rate was established at 300 nL/min. The peptides were scanned by a CaptiveSpray nanolitre ion source into the mass spectrometry, with TIMS turned on in diaPASEF scan mode. The DIA window was delineated in the timsControl program. Each scan cycle included one MS1 scan and a specified diaPASEF scan.

#### Statistical analysis

The DIA raw data files were analyzed using DIA-NN software (version 1.8). The database used for the search was the Proteome Reference Database for Humans in Uniprot. The deep learning technique in DIA-NN predicted a spectrum library, which, together with the spectral library derived from the MBR function, was used to extract DIA raw data for protein quantification information. The final result is screened at 1% FDR for parent ions and protein concentrations. The quantitative proteomic data obtained from screening was used for further analysis.

For functional annotation and enrichment analysis, based on the principle that proteins with the same or similar sequences also have similar functions, the GO term (Biological Process, Cellular Component, Molecular) corresponding to the submitted protein sequences was obtained through the Diamond program of eggNOG-mapper software, KEGG pathway, and COG (Cluster of Orthologous Groups of proteins) categories of the submitted protein sequences. Upon acquiring the annotation data for all proteins identified through mass spectrometry, we extracted pertinent information regarding differentially expressed proteins, quantified the categories and their frequencies, and conducted functional enrichment analysis via hypergeometric testing to identify significant functional categories pertinent to the experiment.

## Results

### Demographic features and clinical characteristics

Due to limited project funding, this study included only 9 plasma samples with well-documented clinical outcomes, which were subjected to repeated proteomic analysis using pooled sample techniques. In postmenopausal obese individuals, ages varied from 42 to 60, with a mean of 53.19; in healthy controls, ages ranged from 41 to 60, with a mean of 52.63. No statistically significant age difference was noticed between the two groups (*P* > 0.05). The initial mean waist circumference for menopausal obesity participants stood at 88cm which decreased to 81cm by 12week following “moxibustion for harmonization of Yin and Yang” treatment. Grouped under moxibustion therapy, the modified Kupperman scores for menopausal obesity individuals averaged 20.21 initially but reduced significantly to 10.37 after twelve weeks compared to their respective baselines(*P* <0 .05), the above results as illustrated in Table1.

### Proteomic profiling of plasma from patients with menopausal obesity

In this project, 9 plasma samples were subjected to mass spectrometry by mixed-sample repetition using DIA proteomics technology. Among them, 3 samples were plasma from menopausal obese patients before moxibustion treatment, 3 samples were plasma from menopausal obese patients after moxibustion treatment, and 3 were plasma from healthy control groups. A total of 630 proteomes and 6314 parent peptides were identified (Figure 2a). Utilizing the previously mentioned quantitative data for each protein, principal component analysis (PCA) and the Pearson correlation coefficient (R) were applied to evaluate the quantitative repeatability of replicated samples and the quantitative correlation among various sample groups. The quantitative protein correlation coefficient distribution heat map (Figure 2b) and the PCA principal component analysis diagram (Figure 2c) reveal that different samples within the same group in this project are clustered within a relatively concentrated range and can be differentiated from the data aggregation area of other groups, indicating that the samples possess favorable quantitative repeatability and correlation, high comparability, and excellent repeatability .

**Figure 2.**
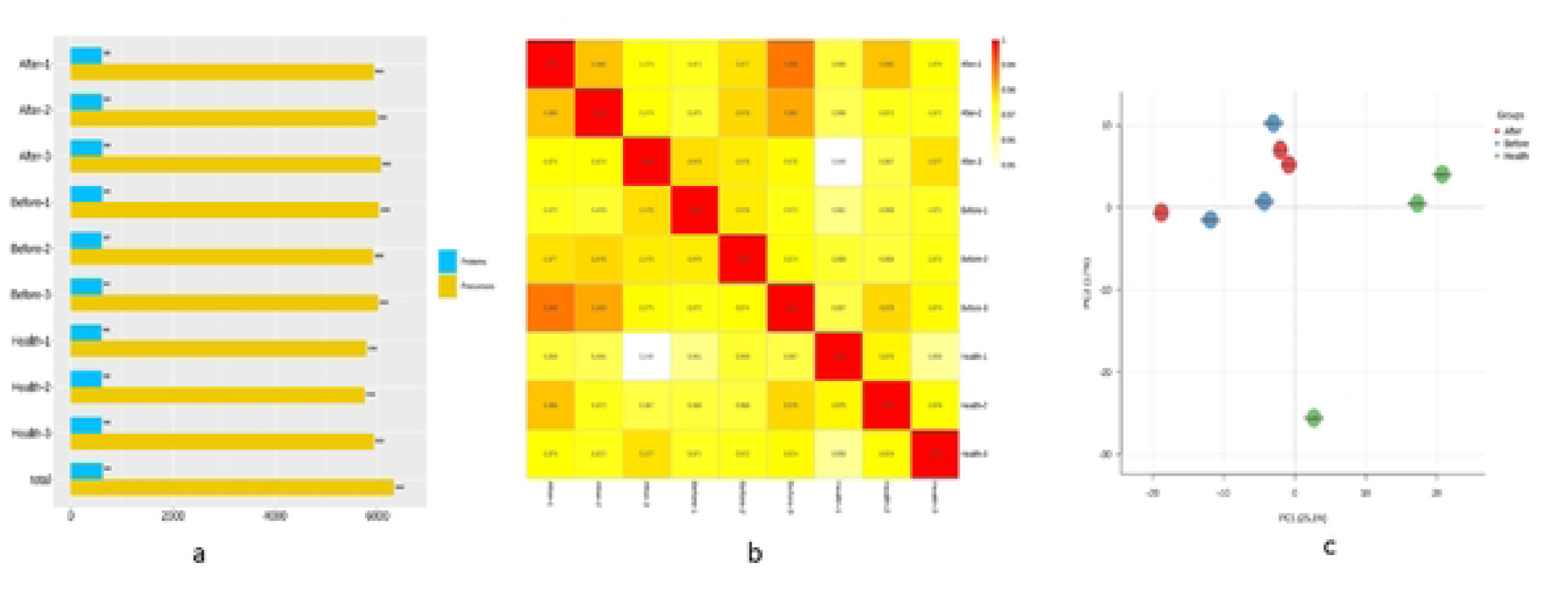
Protein quantification and correlation analysis. a: Proteome and number of parent ions of each sample. b: Quantitative protein correlation coefficient distribution map. c: PCA analysis of differential protein quantification.

### Differentially expressed proteins and bioinformatics analysis between menopausal obese patients and healthy controls

In the menopausal obesity group (Before), 22 proteins exhibited differential expression compared to the healthy control group (Health), with 14 proteins significantly up-regulated and 8 proteins significantly down-regulated, as detailed in Table 2. By annotating and categorising the above differentially expressed proteins in terms of their function, distribution, and pathways involved, the differentially expressed proteins were matched to the classification of each function or pathway. The quantity of differentially expressed proteins matched to each functional or pathway classification and the quantity of markedly enhanced proteins are shown in Figure 3a below.

**Figure 3.**
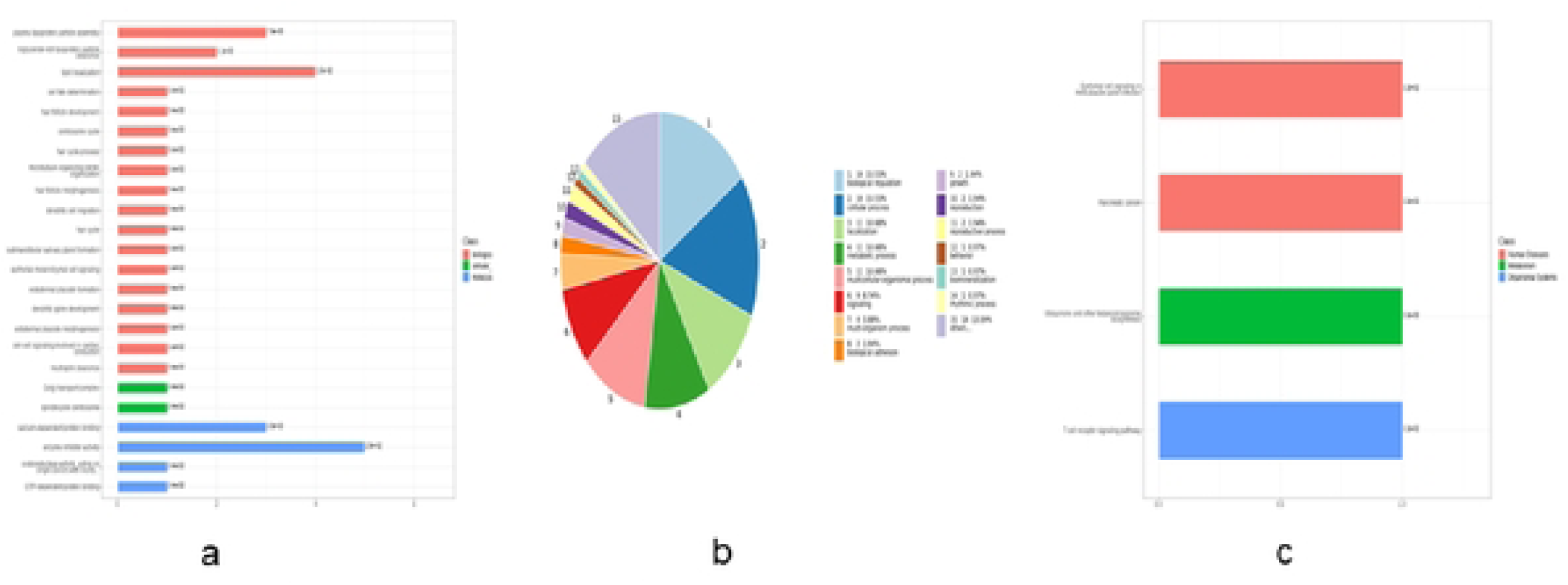
Bioinformatics analysis of differentially expressed proteins in Before/Health group. a: GO enrichment analysis of differentially expressed proteins. b: GO annotation of differential proteins. c: KEGG enrichment of differential proteins.

**Table 1.** Comparison between “Moxibustion for harmonization of Yin and Yang” group(Group A) and Gentle moxibustion group(Group B)

| Variable | Group A | Group B | *P* |
| --- | --- | --- | --- |
| Age (years) | 53.19±4.72 | 52.58±3.92 | 0.624 |
| Disease duration(month) | 12.83±5.14 | 13.15±4.74 | 0.793 |
| Height(cm) | 12.38±40.35 | 6.12±27.48 | 0.270 |
| Weight(kg) | 62.13±69.03 | 63.13±73.18 | <0.05 |
| BMI | 24.38±27.21 | 24.92±28.25 | <0.05 |
| Body fat percentage(%) | 36.97±2.44 | 37.73±3.11 | <0.05 |
| Basal metabolic rate(kcal) | 1439.38±110.04 | 1404.16±124.49 | <0.05 |
| Hip circumference(cm) | 98.49±5.22 | 100.22±5.2 | <0.05 |
| Waist-hip ratio | 0.85±0.91 | 0.86±0.93 | <0.05 |
| waist circumference(cm) | 81±90.38 | 84±94 | <0.05 |
| Kupperman score | 10.37±6.23 | 12.19±4.93 | <0.05 |

Presented data include all participants at baseline and data are presented as mean ± SD or median.

**Table 2.** Differentially expressed proteins in the Before /Health group.

| Protein label | Gene | P-value | Tendency |
| --- | --- | --- | --- |
| A0A0B4J2H0 | IGHV1-69D | 0.021253991 | Up |
| P02655 | APOC2 | 0.018997413 | Up |
| P02656 | APOC3 | 0.017812956 | Up |
| P04083 | ANXA1 | 0.030303348 | Up |
| P20742 | PZP | 0.018040795 | Up |
| P20851 | C4BPB | 0.033906284 | Up |
| P22897 | MRC1 | 0.005334767 | Up |
| P55056 | APOC4 | 0.002613495 | Up |
| P60953 | CDC42 | 0.039168578 | Up |
| Q92954 | PRG4 | 0.001035158 | Up |
| P06748 | NPM1 | 0.024545566 | Up |
| P58166 | INHBE | 0.026990213 | Up |
| P32754 | HPD | 0.024657043 | Up |
| A0A0C4DH39 | IGHV1-58 | 9.95854E-05 | Up |
| A0A075B6I0 | IGLV8-61 | 0.026450206 | Down |
| P11226 | MBL2 | 0.001825787 | Down |
| P24592 | IGFBP6 | 0.030683433 | Down |
| P54289 | CACNA2D1 | 0.014174376 | Down |
| P60985 | KRTDAP | 0.015984021 | Down |
| Q7Z3B1 | NEGR1 | 0.028603622 | Down |
| Q8IUL8 | CILP2 | 0.010985368 | Down |
| P07307 | ASGR2 | 0.001415789 | Down |

The Gene Ontology annotation of differentially expressed proteins in menopausal obesity indicated that these proteins mostly pertained to bioregulatory activities, cellular metabolism, reproductive development, biomineralisation and bioadhesion, and were enriched in the processes of enzyme inhibitor activity, calcium-dependent protein binding, lipid localisation and plasma lipoprotein particle assembly(Figure 3b).

KEGG functional annotation and enrichment study of differentially expressed proteins in menopausal obesity revealed the identification of 11 KEGG pathway genes, with all differentially expressed proteins undergoing KEGG pathway annotation, of which the pathways with p < 0.05 included Epithelial cell signaling in Helicobacter pylori infection, Pancreatic cancer, Ubiquinone and other terpenoid-quinone biosynthesis, and T cell receptor signaling pathway. KEGG annotation entries are shown in Figure 3c.

### Differential expression of proteins and functional enrichment with menopausal obesity patients before and after moxibustion treatment

Three differentially expressed proteins were screened after moxibustion treatment (After) compared with before moxibustion treatment (Before), of which one differently expressed protein was considerably up-regulated, whereas two differentially expressed proteins were significantly down-regulated in Table 3, and the quantitative ratio distribution and the volcano plots of the differentially expressed proteins are shown in Figures 4a and 4b.

**Figure 4.**
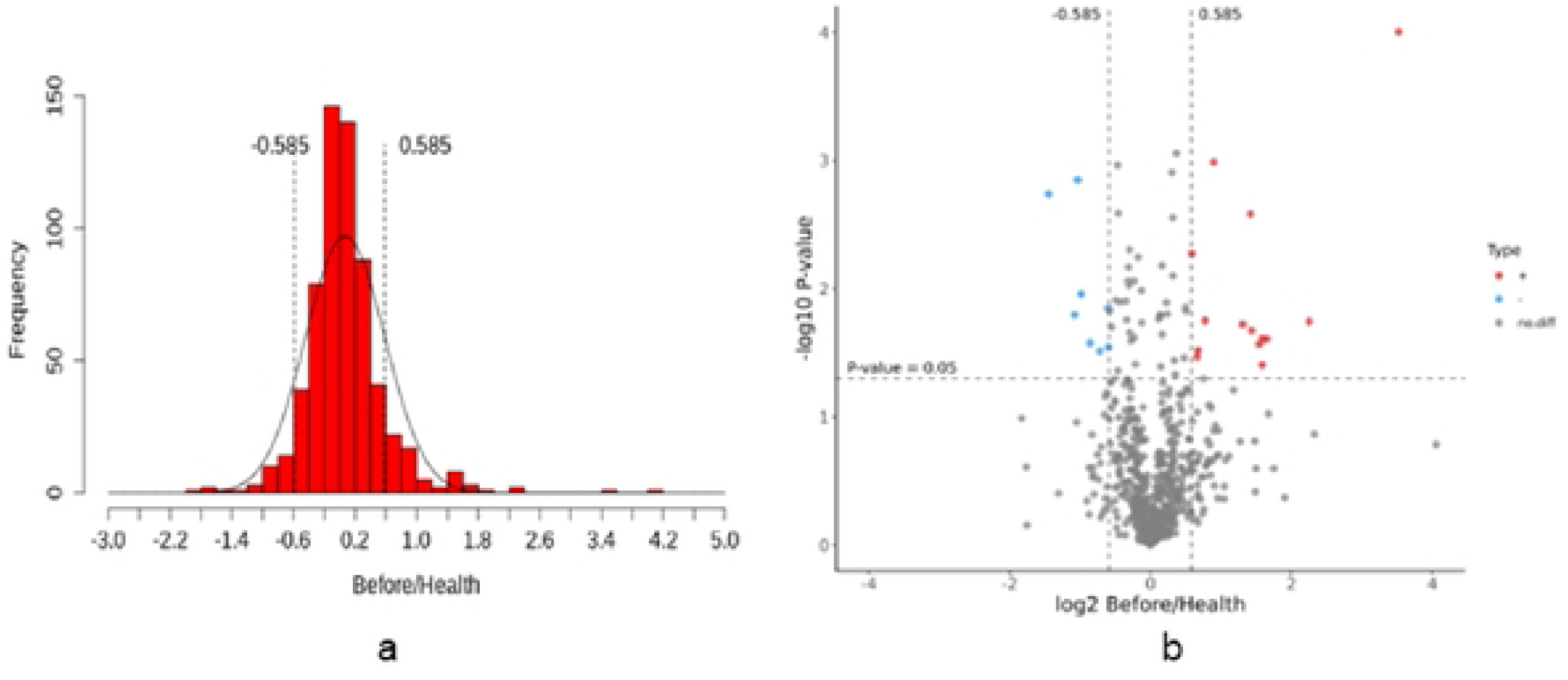
Screening of differentially expressed proteins a: Quantitative ratio distribution of differential proteins. b: Volcano plot of differential proteins.

**Table 3.** Differentially expressed proteins in the After/Before group.

| Protein label | Gene | P-value | Tendency |
| --- | --- | --- | --- |
| P07307 | ASGR2 | 0.015173431 | Down |
| P04083 | ANXA1 | 0.001584671 | Down |
| P07910 | HNRNPC | 0.010266346 | Down |

GO annotation of differentially expressed proteins in menopausal obese patients before and after moxibustion treatment revealed that proteins selectively expressed in menopausal obesity were mainly manifested in the processes of bioregulation, cellular metabolism, growth and development, biomineralisation and immune regulation (Figure 5a), and were enriched in the processes of intracellular phagocytosis, nucleic acid binding, tissue regeneration and neutrophil clearance (Figure 5b).

**Figure 5.**
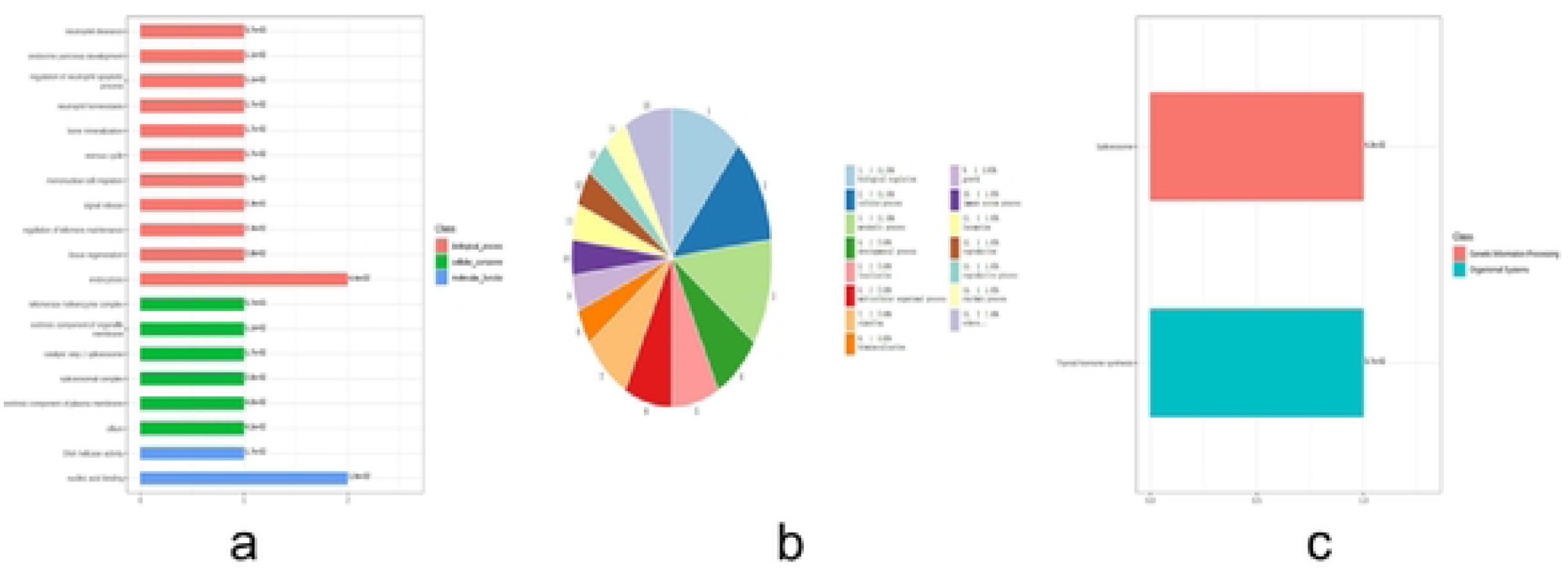
Bioinformatics analysis of differentially expressed proteins in After/Before group. a: GO enrichment analysis of differentially expressed proteins. b: GO annotation of differential proteins. c: KEGG enrichment of differential proteins.

KEGG functional annotation and enrichment analysis of menopausal obese patients before and after moxibustion treatment showed that a total of 2 KEGG pathway genes were annotated (Figure 5c), and all differentially expressed proteins underwent KEGG pathway annotation, of which the pathways with p < 0.05 included Thyroid hormone synthesis and splicing Spliceosome2 signalling pathways

## Discussion

Changes in endogenous sex hormone levels can impact lipid metabolism in adipose tissue of menopausal women. Relative hyperandrogenemia caused by decreased estrogen levels is the primary factor leading to obesity in menopausal women ^[22]^. Relative androgen excess can predict the occurrence of metabolic syndrome during the transition period of menopause, including lipid metabolism disorder and obesity ^[23]^. The increase of body fat in menopausal women mainly occurs centrally, manifested as abdominal fat accumulation. After castration, rats’ exercise amount significantly decreases, resulting in increased adipose tissue and glucose and lipid metabolism disorders ^[24]^. Menopause is a period characterized by increased FSH levels and decreased E2 levels, as well as declining ovarian function.

FSH stimulates hepatic cholesterol production, and serum FSH concentrations correlate favorably with total cholesterol (TC) and low-density lipoprotein cholesterol (LDL-C) levels. FSH enhances the function of the rate-limiting enzyme HMG-CoA in hepatic cholesterol biosynthesis by activating transcription factor Srebp-2, while its action can be blocked by anti-FSHβ antibody or FSHR. This strategy significantly decreases serum and hepatic cholesterol levels in mice ^[25]^. Estradiol (E2) is essential for mitochondrial fatty acid β-oxidation. The interaction between Leptin and Leptin receptor (Lepr) promotes estradiol synthesis, whereas reduced estradiol production inhibits the expression of Leptin and Lepr genes ^[26]^. During menopause, LDL-C cannot be utilized for estrogen synthesis, leading to decreased estrogen production. Postmenopausal model mice exhibit diminished expression of transcription factors required for participation in fatty acid beta oxidation and lipolysis ^[27]^. E2 therapy reinstates mRNA expression and enhances the activity of enzymes associated with fatty acid metabolism in aromatase gene knockout murine models ^[28]^. The reduction of E2 may diminish the expression of genes critical for energy expenditure and those implicated in fatty acid metabolism or lipid catabolism, potentially leading to obesity or metabolic problems in postmenopausal women^[29]^.

This DIA quantitative protein study found significantly higher levels of APOC2 and PZP protein expression in menopausal obese patients than in healthy individuals. APOC2 participates in plasma lipid metabolism and acts as an activator of lipoprotein lipase (LPL). LPL is pivotal in the hydrolysis process of very low density lipoproteins (VLDLs) and coeliac particles ^[30]^ and catalyzes the hydrolysis of triglycerides in plasma lipoproteins on the luminal surface of capillaries which results in the production of tissue-utilised free fatty acids (FFAs) and glycerol, as well as residual lipoproteins that will be cleared ^[31]^.Abnormalities in the APOC2 gene affect LPL activation, and an accumulation of triglyceride-rich chyme and VLDL occurs, which can lead to severe hypertriglyceridaemia.PZP (pregnanza zone protein) is a highly dependent protein in the plasma lipoproteins of the capillary lumen. zone protein) is a highly estrogen-dependent immune plasma protein that can be injected subcutaneously to establish a mouse model of autoimmune ovarian insufficiency ^[32]^. The production of zona pellucida antibodies may be due to the continuous development and atresia of follicles in women of reproductive age, which exposes the body’s immune system to continuous stimulation by zona pellucida antigens, resulting in the production of anti-zona pellucida antibodies, and the high concentration of PZP antibodies causes a decrease in the fertility of the female mice, which affects follicular development and leads to accelerated destruction and depletion of the oocytes, thus causing menopause ^[33]^. Therefore, menopausal obesity is related to the buildup of visceral adipose tissue caused by changes in sex hormone levels and reduced energy consumption after the decline of female ovarian function.

The application of moxibustion to Guanyuan and Sanyinjiao points in perimenopausal model rats can adjust the hormone levels of desiccated rats, resulting in a reduction in FSH levels, an elevation in E2 levels, and therefore an increase in HDL-C, a decrease in TC, TG, and LDL-C, and an improvement in lipid metabolism ^[34]^, which is consistent with the results of our team’s preclinical, large-sample, multicentre, randomised controlled trial ^[35]^. In this study, it was found that after moxibustion treatment, the plasma of menopausal obese patients showed a significant down-regulation of desialylate glycoprotein receptor 2 (ASGR2) protein, membrane-associated protein A1 (ANXA1) and human heterogeneous cytosolic protein (HHPC), and human heterogeneous cytosolic ribonucleoprotein C (HNRNPC), which were enriched in biological processes and molecular functions such as intracellular phagocytosis, nucleic acid binding, tissue regeneration and neutrophil clearance.

Membrane-associated protein A1 (annexin A1, ANXA1) is crucial in regulating obesity and obesity-related metabolic disorders. As an effector of glucocorticoid-mediated responses and a modulator of inflammatory processes, ANXA1 is crucial to innate immune responses, has anti-inflammatory activity and is able to modulate activated T cell activation-triggered signalling cascades by enhancing T cell differentiation and proliferation, contributing to adaptive immune responses and promoting inflammatory absorption and wound healing ^[36]^. Studies have shown ^[37]^ that ANXA1 competitively binds to PDZ and LIM structural domain 7 (PDLIM7) via MYC binding protein 2 (MYCBP2), which in turn affects SMAD4 ubiquitylated proteasomal degradation and adipogenic processes; Examination of subcutaneous adipose tissue (SAT) in humans and mice showed that ANXA1 was abnormally elevated in SAT of the obese group and that ANXA1 protein expression was significantly decreased in plasma of menopausal obese patients after moxibustion treatment.

HNRNPC is a crucial RNA-binding protein, extensively expressed in cells, that facilitates intron removal and the accurate assembly of exons during RNA splicing ^[38]^. The calcium-sensitive receptor (CaSR) is a crucial membrane protein that is part of the G protein-coupled receptor family C. CaSR agonists cause a significant increase in AKT activity, and its inhibitors inhibit AKT activity. Using gene microarray technology, it was found that in the liver of high-fat diet-induced insulin resistance (IR) rats, HNRNPC protein expression was significantly increased, CaSR expression was downregulated, and the level of Akt phosphorylation in the IR-associated signalling pathway PI3K/Akt was reduced, leading to IR ^[39]^. After moxibustion treatment, HNRNPC expression was significantly decreased, suggesting that moxibustion can ameliorate insulin resistance.

ASGR2 is a protein-coding gene whose protein product is involved in protein metabolism and transport to the Golgi and subsequent modification ^[40]^. Receiver operating characteristic (ROC) curves and nomogram analyses of microarray data from patients with myocardial infarction (MI) at 1 month and 6 months post-discharge showed that ASGR2 expression was markedly elevated compared to that of control individuals without a history of MI, and the results of Pearson correlation analysis showed that the ASGR2 gene was highly correlated with most of the iron death-related genes, which may lead to increased myocardial remodelling and more severe dysfunction after myocardial infarction by triggering inflammatory responses ^[41]^. ASGR is predominantly expressed in liver parenchymal cells, and ASGR2 is capable of extracellular secretion.As a lectin, it recognises and takes up glycoproteins containing N-acetylgalactosamine or galactose residues and removes desialylated glycoproteins from the blood.ASGR has an affinity for LDL and chymotrypsin and may play a role in the removal of chymotrypsin from the blood. In a mouse model,Ldlr knockout,Asgr2 knockout,LDL,TC and TG in the blood were reduced to different degrees.Asgr2 deletion can improve the metabolic capacity of the body and alleviate metabolic syndrome caused by high-fat diet^[42]^. After moxibustion treatment, the expression of ASGR2 was significantly reduced, which not only improved abnormalities of glucose and lipid metabolism, but also reduced the risk of cardiovascular and cerebrovascular diseases caused by menopausal obesity.

Autophagy removes damaged and exhausted organelles, enables energy recycling and reuse, and slows down cellular aging. Autophagy is closely associated with follicular reserve, follicular development and atresia ^[43]^, and is a key factor in maintaining ovarian follicular viability. The PI3K/Akt/mTOR pathway is a key signalling pathway for cellular autophagy regulation and resistance to apoptosis, and activation of the PI3K/Akt/mTOR autophagy signalling pathway not only delays ovarian aging ^[44]^, but also reduces obesity and insulin resistance ^[45]^. After moxibustion intervention, the biological processes and molecular functions of significantly differentially expressed proteins were enriched in endocytosis, nucleic acid binding, tissue regeneration and neutrophil clearance in menopausal obese patients. Therefore, we hypothesize that the mechanism by which moxibustion addresses menopausal obesity may include the downregulation of HNRNPC expression, activation of the PI3K/Akt/mTOR autophagy signalling pathway, regulation of hormone levels, and consequently delay of ovarian aging and improvement of lipid metabolism. This study has elucidated the pathogenesis of menopausal obesity, which provides ideas and clues for the mechanism of moxibustion in the management of menopausal obesity, but how moxibustion regulates the upstream and downstream factors of the PI3K/Akt/mTOR pathway needs to be verified by further in-depth animal experiments.

## Data Availability

Data available on request from the authors.

## Ethics approval

The study design complies with the guidelines of the Declaration of Helsinki (Version Edinburgh 2000), meets medical ethics Requirements and has been approved by the Ethics Committee of Hubei Provincial Hospital of Traditional Chinese Medicine (HBZY2021-C17-01).

## Funding

The trial is financially supported by the National Administration of Traditional Chinese Medicine (No.0686-204001171345N, GZY-KJS-2020-078), Qhuang Project of the National Administration of Traditional Chinese Medicine (No.[2018]284), the 2nd Hubei Province’s Outstanding Medical Academic Leader Program (Health Commission of Hubei Province Notice No.[2019]47) and the first Tanhualin famous doctor, student training project (Hubei Traditional Chinese Medicine Student: No.[2018]72). The sponsor played no part in the study design; collection, management, analysis, and interpretation of data; writing of the report; and the decision to submit the report for publication.

## Informed Consent Statement

Detailed information of the trial was informed to the participants, and written informed consent was obtained from every participant before screening.

## Disclosure

The funders had no role in the design, execution, or writing of the study.

## Conflicts of Interest

The authors declare that they have no competing interests.

## Authors’ contributions

BLY and YBZ contributed equally to the study, ZYZ conceived and designed the study. ZYZ take responsibility for the integrity of the data and the accuracy of the data analysis. BLY and YBZ drafted the manuscript. ZYZ revised the manuscript. YBZ recruited, treated and followed up patients. All authors reviewed and approved the final version of the manuscript.

Benlu Yu and Yunbao Zhu contributed equally to this work. Conceptualization: Benlu Yu. Data curation: Benlu Yu. Formal analysis: Yunbao Zhu. Funding acquisition: Zhongyu Zhou. Investigation: Benlu Yu. Methodology: Benlu Yu. Project administration: Zhongyu Zhou. Resources: Zhongyu Zhou. Supervision: Benlu Yu,. Validation: Yunbao Zhu. Visualization: Yunbao Zhu. Writing-original draft: Benlu Yu, Yunbao Zhu. Writing-review & editing:Zhongyu Zhou. All authors reviewed and approved the final version of the manuscript.

## Notes

### Competing Interest Statement

The authors have declared no competing interest.

### Clinical Trial

NCT04943705

### Funding Statement

Yes

### Author Declarations

Ethics Committee of Hubei Provincial Hospital of Traditional Chinese Medicine

